# Nocturnal Respiratory Rate and Variability Predict Long-term Mortality in Stable Outpatients with Cardiovascular Disease

**DOI:** 10.64898/2026.06.12.26355214

**Authors:** Raimon Padrós-Valls, Keshav Gupta, Nick Harrington, Justin Yu, Jeremy E. Orr, Robert L. Owens, David Torres Barba, Kevin R. King

## Abstract

**Background:** Respiratory rate (RR) predicts short-term mortality in acute care settings, yet its prognostic significance in clinically stable outpatients remains poorly defined.

**Objectives:** To determine whether the median and variability of nocturnal respiratory rate (NRR) are independently associated with long-term cardiovascular and all-cause mortality in outpatients with cardiovascular disease.

**Methods:** We analyzed overnight chest belt waveforms from elective polysomnography in 5,679 older adults with cardiovascular disease enrolled in the Sleep Heart Health Study (SHHS). NRR was quantified at 30-second resolution, and per-subject median NRR and within-night variability (standard deviation) were derived. Kaplan-Meier survival analysis and Cox proportional hazards models were used to evaluate associations with cardiovascular and all-cause mortality over 3-year and 15-year follow-up periods, adjusting for demographic characteristics, cardiopulmonary comorbidities, and sleep apnea severity.

**Results:** Higher median NRR and greater NRR variability were each associated with increased cardiovascular and all-cause mortality. Combining these metrics identified a high-risk group characterized by high median and high variability of NRR, with nearly five-fold higher 3-year all-cause mortality compared with a low-risk group (unadjusted HR: 2.61; 95% CI: 1.65, 4.14; *p*<0.001; adjusted HR: 2.15; 95% CI: 1.30, 3.55; *p*=0.003).

**Conclusions:** Both the baseline level and variability of NRR independently predict morttality in clinically stable outpatients with cardiovascular disease. Densely profiled NRR represents a promising, underutilized biomarker for long-term risk stratification.

**Condensed Abstract:** Nocturnal respiratory rate (NRR) is an underutilized biomarker whose prognostic significance in stable cardiovascular outpatients is unknown. In 5,679 participants from the Sleep Heart Health Study, median NRR and within-night variability derived from overnight polysomnography independently predicted cardiovascular and all-cause mortality. Stratification based on these metrics identified a high-risk group with nearly five-fold higher 3-year mortality compared with a low-risk group (adjusted HR: 2.15; 95% CI: 1.30-3.55; *p*=0.003).

## Introduction

Respiratory rate (RR) is a fundamental vital sign, yet it remains one of the least precisely measured and least utilized in clinical practice despite evidence that deviations in RR often precede clinical deterioration and predict mortality across inpatient settings^1,2^. RR is inconsistently documented in routine care and suffers from measurement error due to infrequent manual sampling, and susceptibility to confounding by activity, emotion, and observer variability^3^. This has limited its clinical utility outside of extreme physiological states. Nonetheless, elevated RR is a strong predictor of short-term mortality, intensive care transfer, and clinical deterioration and is incorporated into early warning systems where the respiratory component often contributes significantly to discrimination of acute risk^4–13^.

Nocturnal respiratory rate (NRR) – the frequency of breathing during sleep – offers a complementary and potentially more informative physiologic signal. During sleep, respiratory control is relatively free from behavioral and emotional confounders, enabling assessment of fundamental cardiopulmonary and autonomic physiology^14–16^. Advances in wearable and non-contact sensing technologies have made longitudinal automated measurement of NRR increasingly feasible in both inpatients and outpatient settings^17^. Importantly, raw respiratory waveforms collected during routine overnight polysomnography (PSG) provide an existing, underexploited resource for characterizing NRR dynamics at high temporal resolution^12^.

Prior studies linking respiratory rate to adverse outcomes have largely focused on acute or sparsely sampled awake measurements, such as respiratory rate recorded during or shortly after acute myocardial infarction or acute coronary syndromes, where elevations likely reflect transient physiological stress rather than stable respiratory control mechanisms^8,13^. More recently, analyses of overnight polysomnography in community-based cohorts have shown that higher mean nocturnal respiratory rate (NRR) is associated with increased long-term mortality^12^. However, these studies were conducted primarily in older adults without a specific focus on established cardiovascular disease and relied on single summary measures of NRR, without evaluating within-night variability of respiratory rate as a distinct physiological signal.

At the same time, contemporary implantable cardiac devices incorporate nighttime respiratory metrics as key inputs to multiparameter risk algorithms for heart failure decompensation, reflecting sensitivity of nocturnal respiratory patterns to cardiopulmonary load, fluid status, and autonomic regulation^18–21^. Yet, the independent prognostic contribution of respiratory rate within these proprietary systems remains opaque, and their applicability is limited to selected populations with advanced disease and implanted hardware. Consequently, despite growing recognition that nocturnal respiratory physiology carries clinically relevant information, the prognostic significance of both the baseline level and variability of NRR in clinically stable outpatients with cardiovascular disease remains poorly defined.

In this study, we address these gaps by leveraging waveform-level analysis of overnight polysomnography in a large, well-characterized cohort of stable outpatients with cardiovascular disease. By jointly examining the median and within-night variability of nocturnal respiratory rate, we test the hypothesis that both baseline respiratory control and its short-term instability carry prognostic information, and that their combination enables improved mortality risk stratification beyond prior approaches. We find that higher median NRR and greater NRR variability were each associated with increased cardiovascular and all-cause mortality. Combining these metrics identified a high-risk group with elevated median NRR and NRR variability that had nearly five-fold higher 3-year all-cause mortality compared with the low-risk group.

## Methods

### Study Design and Population

The SHHS is a multi-center prospective cohort study of the cardiovascular and other health consequences of sleep-disordered breathing (SDB). The study design, sampling methods, and protocols have been reported previously^22^. The cohort is composed of participants from existing NHLBI epidemiological studies including Framingham Offspring Cohort, Atherosclerosis Risk in Communities Study (ARIC), Cardiovascular Health Study (CHS), Strong Heart Study (SHS), the New York Hypertension Cohorts, the Tucson Epidemiologic Study of Airways Obstructive Diseases and the Health and Environment Study. The dataset includes participants at least 40 years of age who lacked a history of sleep apnea treatments or tracheostomies, and who were not undergoing home oxygen therapy. Some cohorts oversampled for snorers younger than 65 years of age to increase the prevalence of SDBs to produce a cohort of 6,441 participants, though due to sovereignty issues, the SHS participants were excluded, yielding a final shared dataset of 5,802 patients. Participants provided written consent and participating institution site institutional review boards approved the study protocols. The final cohort with median NRR available was 5,679 patients^23^. The data was accessed through an agreement with the National Sleep Research Resource. A flowchart describing how the study cohort was derived is shown in **Figure 1**, and a summary of the cohort demographics and characteristics is provided in **Table 1**.

**Figure 1.**
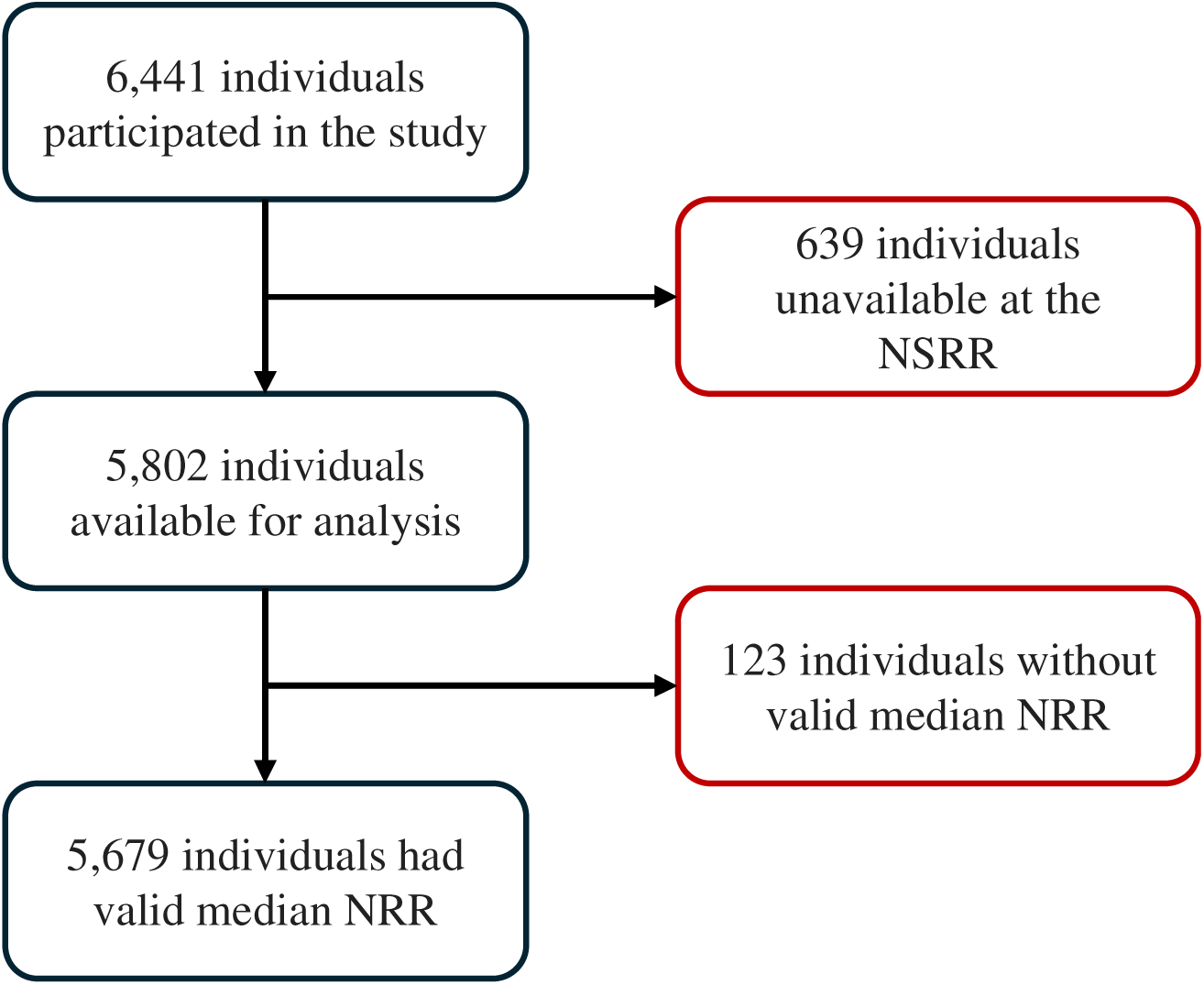
Study cohort derivation. Of the original SHHS cohort (n=6,441), we excluded individuals without polysomnography data available in the NSRR database (n=639), and those individuals without a valid median nocturnal respiratory rate measurement (n=123; see *Methods* section for more details), resulting in a final study cohort of 5,679 participants.

**Table 1.** SHHS cohort demographics, baseline characteristics, and outcome incidence at study enrollment. Continuous variables are reported as *mean (standard deviation)*, while categorical and dichotomous variables are presented as *count (%)*.

|  | N | 3-Year Horizon |  |  | 15-Year Horizon |  |  |
| --- | --- | --- | --- | --- | --- | --- | --- |
|  |  | Mortality | Survival | P-Value | Mortality | Survival | P-Value |
| Overall | 5,679 | 190 (3.35) | 5,489 (96.65) |  | 1,276 (22.47) | 4,403 (77.53) |  |
| Censoring Date (years) | 11.00 (3.16) | 1.76 (0.83) | 2.99 (0.19) | **<0.001** | 7.03 (3.36) | 12.15 (1.93) | **<0.001** |
| Median NRR (brpm) | 15.22 (2.37) | 16.66 (3.05) | 15.17 (2.32) | **<0.001** | 15.87 (2.64) | 15.03 (2.24) | **<0.001** |
| SD NRR (brpm) | 1.10 (0.42) | 1.26 (0.56) | 1.09 (0.42) | **<0.001** | 1.14 (0.49) | 1.08 (0.4) | **<0.001** |
| SBP (mmHg) | 127.34 (19.33) | 132.16 (20.14) | 127.17 (19.28) | **<0.001** | 132.65 (20.37) | 125.78 (18.73) | **<0.001** |
| DBP (mmHg) | 73.64 (11.63) | 70.02 (12.71) | 73.77 (11.57) | **<0.001** | 70.61 (12.48) | 74.53 (11.21) | **<0.001** |
| Age (years) | 63.15 (11.24) | 74.36 (10.18) | 62.76 (11.08) | **<0.001** | 72.63 (9.31) | 60.4 (10.22) | **<0.001** |
| BMI (kg/m<sup>2</sup>) | 28.16 (5.09) | 27.26 (5.01) | 28.19 (5.09) | **0.013** | 27.7 (5.05) | 28.29 (5.1) | **<0.001** |
| Female | 2977 (52.42) | 81 (42.63) | 2,896 (52.76) | **0.007** | 590 (46.24) | 2,387 (54.21) | **<0.001** |
| Race |  |  |  | **0.028** |  |  | **<0.001** |
| White | 4801 (84.54) | 165 (86.84) | 4,636 (84.46) |  | 1,107 (86.76) | 3,694 (83.9) |  |
| Black | 505 (8.89) | 21 (11.05) | 484 (8.82) |  | 140 (10.97) | 365 (8.29) |  |
| Other | 373 (6.57) | 4 (2.11) | 369 (6.72) |  | 29 (2.27) | 344 (7.81) |  |
| Hispanic or Latino | 272 (4.79) | 2 (1.05) | 270 (4.92) | **0.023** | 19 (1.49) | 253 (5.75) | **<0.001** |
| Heart Failure | 96 (1.77) | 20 (10.93) | 76 (1.45) | **<0.001** | 67 (5.46) | 29 (0.69) | **<0.001** |
| Atrial Fibrillation | 62 (2.12) | 10 (7.41) | 52 (1.87) | **<0.001** | 48 (5.52) | 14 (0.68) | **<0.001** |
| Diabetes | 400 (7.38) | 40 (21.28) | 360 (6.88) | **<0.001** | 194 (15.63) | 206 (4.93) | **<0.001** |
| Hypertension | 2,421 (42.63) | 118 (62.11) | 2,303 (41.96) | **<0.001** | 790 (61.91) | 1,631 (37.04) | **<0.001** |
| LBBB | 44 (1.09) | 5 (3.55) | 39 (1.0) | **0.017** | 26 (2.66) | 18 (0.59) | **<0.001** |
| RBBB | 110 (2.72) | 12 (8.51) | 98 (2.51) | **<0.001** | 50 (5.12) | 60 (1.96) | **<0.001** |
| History of MI | 337 (6.82) | 30 (17.65) | 307 (6.43) | **<0.001** | 174 (15.12) | 163 (4.3) | **<0.001** |
| CVD Death | 351 (7.1) | 55 (32.35) | 296 (6.2) |  | 351 (30.5) |  |  |
| COPD | 62 (1.11) | 4 (2.19) | 58 (1.07) | 0.145 | 27 (2.17) | 35 (0.81) | **<0.001** |
| Smoke | 2,987 (52.94) | 111 (58.42) | 2,876 (52.75) | 0.143 | 749 (58.88) | 2,238 (51.21) | **<0.001** |
| Asthma | 489 (8.75) | 18 (9.57) | 471 (8.72) | 0.783 | 100 (7.97) | 389 (8.98) | 0.290 |
| AHI (events/hour) | 8.58 (12.21) | 11.53 (15.32) | 8.48 (12.08) | **0.007** | 10.32 (13.23) | 8.08 (11.85) | **<0.001** |

AHI: Apnea-Hypopnea Index; BMI: Body Mass Index; brpm: breaths per minute; COPD: Chronic Obstructive Pulmonary Disease; CVD: Cardiovascular Disease; DBP: Diastolic Blood Pressure; LBBB: Left Bundle Branch Block; MI: Myocardial Infarction; NRR: Nocturnal Respiratory Rate; RBBB: Right Bundle Branch Block; SBP: Systolic Blood Pressure; SD: Standard Deviation.

### Quantification of Respiratory Rate

For each participant’s first sleep study, the chest respirometer waveform was used to estimate the median and standard deviation (SD) of NRR of the individual. The raw signal, sampled at 10 Hz, was first processed using a low-pass filter with a 0.5 Hz cutoff frequency. Peak detection was performed to the resulting signal to determine timepoints when an individual was taking a breath. A sample night with three different 1-minute windows illustrating the results of the peak detection algorithm is depicted in **Online Figure 1**.

Instantaneous values of RR were computed using a sliding window approach (window size of 5 minutes with 1-minute stride). For every window, the RR was calculated by dividing the number of complete breaths (number of peaks minus one) by the total duration of these breaths (time between the first and last peak in seconds) and then converting this value to breaths per minute, which can be described by the formula: 60 * (# peaks -1) / time between first and last peak.

To reduce noise, only 5-minute windows in which all corresponding intrabreath intervals values fell within a physiologically reasonable range (5 to 50 breaths per minute (brpm)) were included in the analysis. To further ensure data quality, the first and last hour of recording were excluded to remove periods of sleep onset and waking. Participants with fewer than one hour of valid RR data (123 individuals) were excluded from the final dataset.

Finally, to compare baseline NRR, we calculated the median of the valid RR windows, while within-night variability was assessed using the standard deviation across RR windows. A summary of the data curation process is provided in **Online Table 1**.

### Statistical Analyses

In **Table 1**, the group of individuals who died was compared to those who survived (in a 3-year follow-up and 15-year follow-up period) using a Student’s t-test or Kruskal-Wallis test as appropriate for continuous variables and a Chi-Square test or Fisher’s exact test as appropriate for dichotomous and categorical variables.

Median NRR was binned for Kaplan-Meier survival analysis every 2 breaths per minute (brpm). Similarly, SD NRR was binned every 0.25 brpm. Statistical differences between survival curves were assessed using the log-rank test.

Risk groups were derived using quintiles. The fifth quintile was used to define the population with high median NRR and high SD NRR separately. The fifth quintiles were 17 and 1.35 brpm for median and SD NRR respectively. Four groups were derived based on these thresholds: high median and SD NRR (High-risk), low median and SD NRR (Low-risk), low median NRR and high SD NRR (Mid-risk 1) and high median NRR and low SD NRR (Mid-risk 2). Kaplan-Meier Survival curves for the two mid-risk groups showed an almost identical trajectory, warranting their combination into a unique mid-risk group. Risk groups characteristics were compared in **Table 2** using the same approach as in **Table 1**.

**Table 2.** NRR based risk groups demographics, baseline characteristics, and outcome incidence at study enrollment. Continuous variables are reported as *mean (standard deviation)*, while categorical and dichotomous variables are presented as *count (%)*.

|  | **N** | **Low Risk** | **Mid Risk** | **High Risk** | **P-Value** |
| --- | --- | --- | --- | --- | --- |
| Overall | 5,679 | 3828 (67.41) | 1439 (25.34) | 412 (7.25) |  |
| Death | 1276 (22.47) | 730 (19.07) | 400 (27.8) | 146 (35.44) | **<0.001** |
| CVD Death | 351 (7.1) | 205 (6.11) | 108 (8.77) | 38 (10.67) | **<0.001** |
| Censoring Date (years) | 11.0 (3.16) | 11.22 (2.88) | 10.71 (3.45) | 9.89 (4.08) | **<0.001** |
| Median NRR (brpm) | 15.22 (2.37) | 14.3 (1.55) | 16.47 (2.36) | 19.41 (2.1) | **<0.001** |
| SD NRR (brpm) | 1.1 (0.42) | 0.92 (0.21) | 1.36 (0.48) | 1.78 (0.5) | **<0.001** |
| SBP (mmHg) | 127.34 (19.33) | 127.02 (19.29) | 127.22 (18.84) | 130.76 (20.98) | **0.001** |
| DBP (mmHg) | 73.64 (11.63) | 73.59 (11.42) | 73.45 (11.71) | 74.83 (13.06) | 0.098 |
| Age (years) | 63.15 (11.24) | 62.79 (10.98) | 63.74 (11.63) | 64.37 (12.1) | **0.002** |
| BMI (kg/m<sup>2</sup>) | 28.16 (5.09) | 28.03 (4.95) | 28.16 (5.14) | 29.31 (6.04) | **<0.001** |
| Female | 2977 (52.42) | 2064 (53.92) | 711 (49.41) | 202 (49.03) | **0.005** |
| Race |  |  |  |  | **<0.001** |
| White | 4801 (84.54) | 3283 (85.76) | 1195 (83.04) | 323 (78.4) |  |
| Black | 505 (8.89) | 299 (7.81) | 155 (10.77) | 51 (12.38) |  |
| Other | 373 (6.57) | 246 (6.43) | 89 (6.18) | 38 (9.22) |  |
| Hispanic or Latino | 272 (4.79) | 178 (4.65) | 63 (4.38) | 31 (7.52) | **0.024** |
| Heart Failure | 96 (1.77) | 41 (1.12) | 33 (2.42) | 22 (5.57) | **<0.001** |
| Atrial Fibrillation | 62 (2.12) | 28 (1.45) | 22 (2.93) | 12 (5.08) | **<0.001** |
| Diabetes | 400 (7.38) | 224 (6.13) | 128 (9.3) | 48 (12.53) | **<0.001** |
| Hypertension | 2,421 (42.63) | 1548 (40.44) | 652 (45.31) | 221 (53.64) | **<0.001** |
| LBBB | 44 (1.09) | 26 (0.94) | 11 (1.11) | 7 (2.4) | 0.074 |
| RBBB | 110 (2.72) | 77 (2.79) | 25 (2.52) | 8 (2.74) | 0.899 |
| History of MI | 337 (6.82) | 186 (5.54) | 105 (8.52) | 46 (12.92) | **<0.001** |
| COPD | 62 (1.11) | 29 (0.77) | 21 (1.5) | 12 (2.98) | **<0.001** |
| Smoke | 2,987 (52.94) | 1930 (50.68) | 810 (56.84) | 247 (60.39) | **<0.001** |
| Asthma | 489 (8.75) | 304 (8.07) | 130 (9.19) | 55 (13.55) | **<0.001** |
| AHI (events/hour) | 8.58 (12.21) | 8.04 (11.37) | 9.04 (12.3) | 12.01 (17.65) | **<0.001** |

AHI: Apnea-Hypopnea Index; BMI: Body Mass Index; brpm: breaths per minute; COPD: Chronic Obstructive Pulmonary Disease; CVD: Cardiovascular Disease; DBP: Diastolic Blood Pressure; LBBB: Left Bundle Branch Block; MI: Myocardial Infarction; NRR: Nocturnal
Respiratory Rate; RBBB: Right Bundle Branch Block; SBP: Systolic Blood Pressure; SD: Standard Deviation.

Univariate and multivariate Cox proportional-hazard models were constructed to assess the mortality hazard ratio of the different risk groups when controlling for different comorbidities including age, gender, obesity (BMI>30), heart failure, atrial fibrillation, diabetes, hypertension, COPD, asthma, history of smoking, and severe sleep apnea (apnea-hypopnea index > 30). The hazard ratio, together with the 95% confidence interval and the corresponding p-value are reported. The Schoenfeld residuals were used to evaluate whether the proportional hazards assumption was met.

Significance was set at p < 0.05 for all the analyses. Data preprocessing and statistical analysis was performed in Python using open-source libraries including scikit-learn, NumPy, SciPy, Pandas, lifelines, and statsmodels.

## Results

### Participant Characteristics

Respiratory waveforms are routinely collected during overnight PSGs using a respiratory inductance plethysmograph, or chest belt. Since RR or NRR metrics are not routinely quantified as part of PSGs, we developed, validated, and applied a waveform-level analysis algorithm based on peak-finding and time-averaged interbreath interval quantification to remove low quality signals and quantify time-averaged NRR at 30-second intervals. Of the 5,802 SHHS PSG participants, 123 studies were excluded due to lack of sufficient high quality chest belt waveform data (**Figure 1**). The resulting final cohort (n=5,679) was gender-balanced, 2,702 (47.58%) men and 2,977 (52.42%) women, had a mean age of 63.2 (SD 11.2) years and a mean BMI of 29.16 (SD 5.09). The subjects had a range of cardiopulmonary and systemic comorbidities with potential to influence NRR including 52.94% with a history of smoking, 42.63% with hypertension, 7.38% with diabetes, 2.12% with atrial fibrillation, 1.77% with heart failure, and 1.11% with chronic obstructive pulmonary disease (COPD) (**Table 1**).

### Nocturnal Respiratory Rate Distributions

The distributions of NRR median and NRR SD are shown in **Figure 2**. The median follows a distribution centered around 15 brpm, with a small rightward skew. In contrast, the distribution of SDs of NRR was centered around 1.1 brpm and exhibited a strong rightward skew.

**Figure 2.**
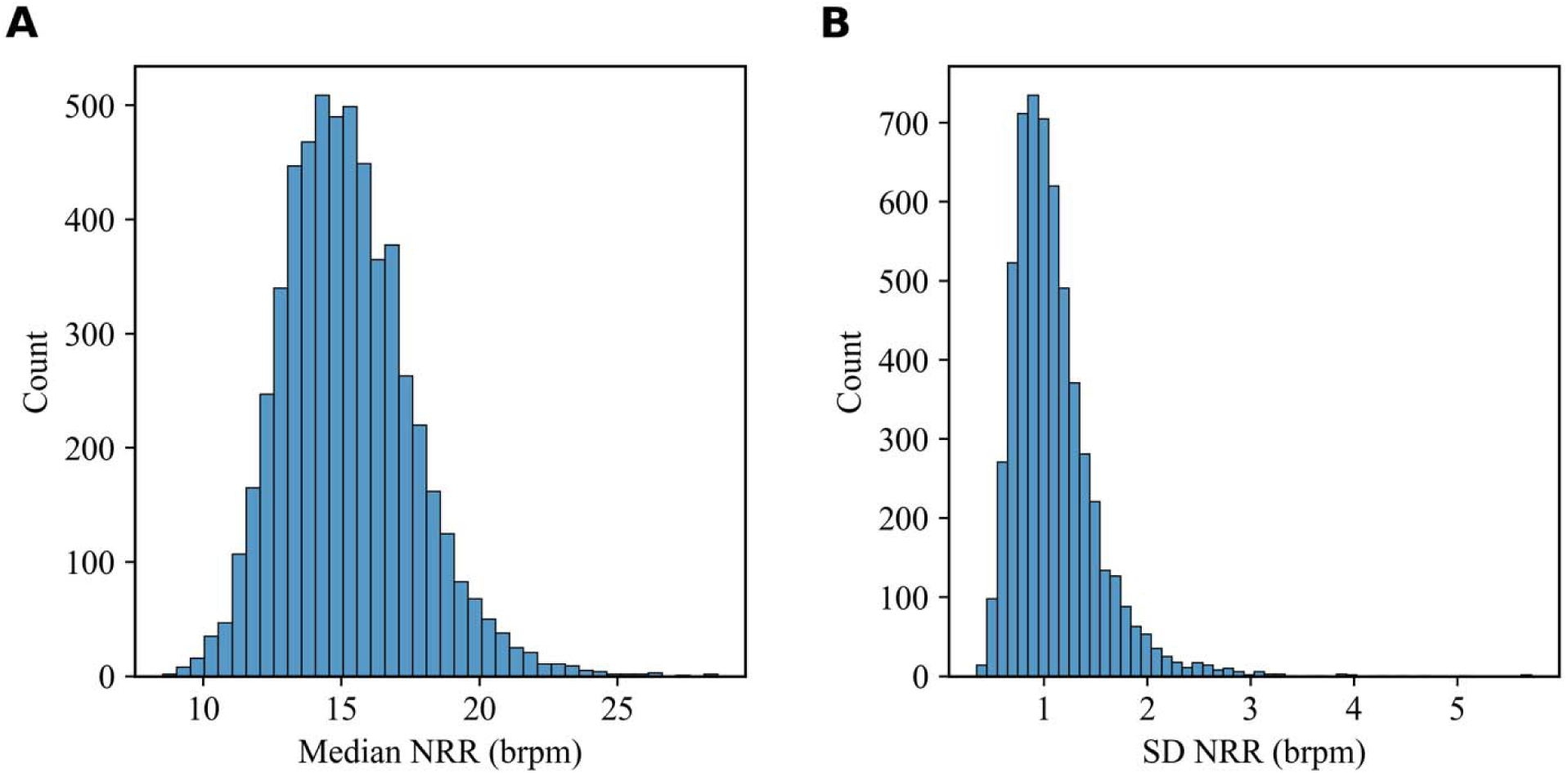
Distribution of NRR derived biomarkers. (A) Histogram of median NRR. (B) Histogram of NRR standard deviation. n=5,679. brpm: breaths per minute; NRR: nocturnal respiratory rate; SD: standard deviation.

### NRR-based Survival – Univariate Analysis

We first analyzed the relationship between median NRRs and mortality. Kaplan-Meier curves revealed a significant dose-dependent relationship between median NRR and cardiovascular and all-cause mortality. Higher median NRR was associated with a significant increased risk of mortality over both 3-year and 15-year follow-up period (**Figure 3A, C; Online Figures 2A, C; 3A, C; and 4A, C**). P-values obtained from a log-rank test comparing each pair of curves are provided in **Online Table 2**. We next performed a similar analysis based on the variability of NRR during each night by relating SD of NRR to cardiovascular and all-cause mortality (**Figure 3B, D; Online Figures 2B, D; 3B, D; and 4B, D**). In both cases, individuals with high NRR SD exhibited significantly lower survival compared to individuals with low NRR SD. The p-value obtained from a log-rank test comparing each pair of curves are provided in **Online Table 3**.

**Figure 3.**
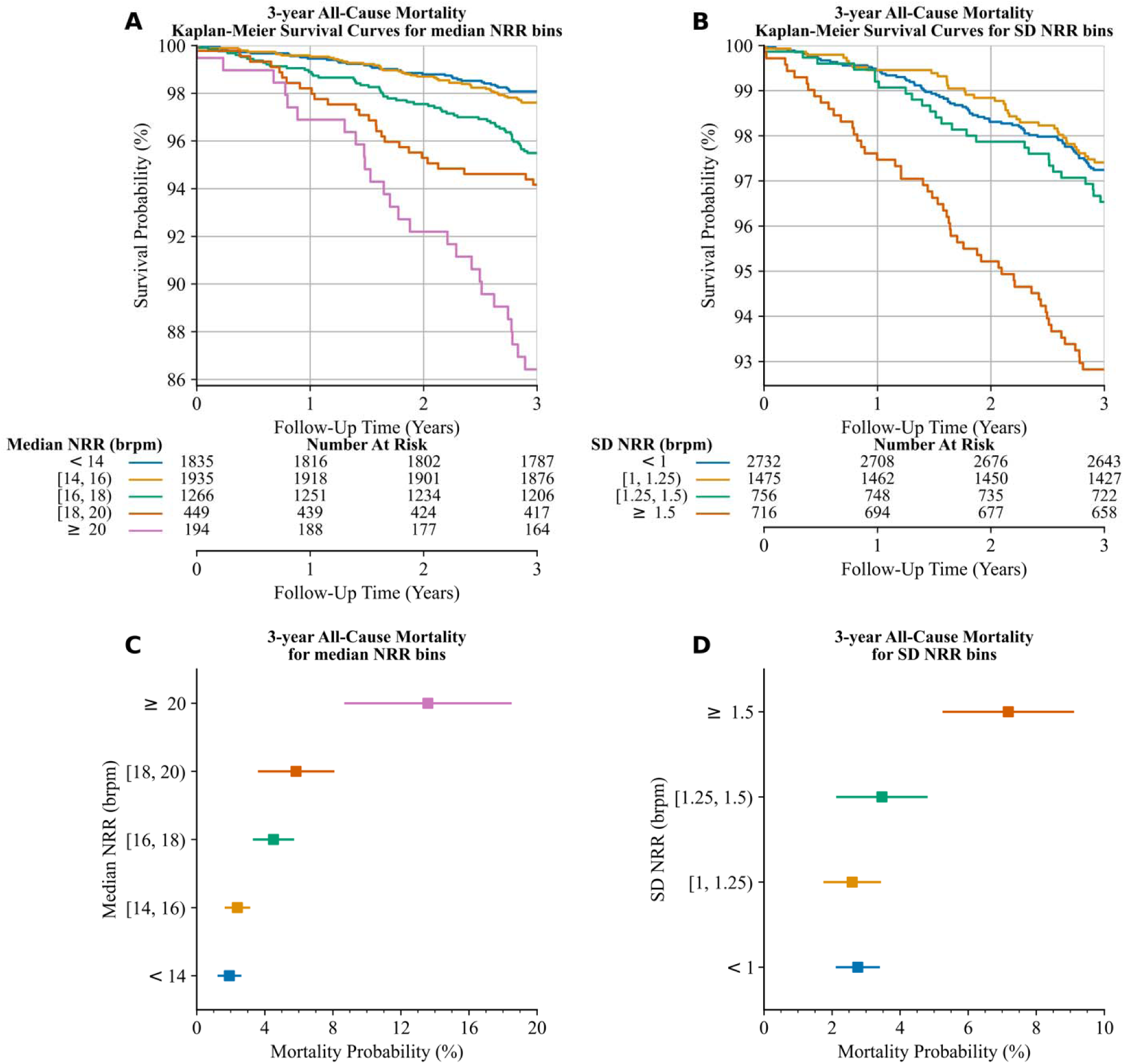
3-year all-cause mortality analysis for bins of median NRR and bins of SD NRR. Kaplan-Meier survival curves in a 3-year horizon (A) for median NRR bins, and (B) for SD NRR bins. 3-year cumulative incidence of death (C) for median NRR bins, and (D) for SD NRR bins. brpm: breaths per minute; NRR: nocturnal respiratory rate; SD: standard deviation.

### NRR-based Risk Stratification

Survival curves based on NRR medians and SDs were used to define 3 risk groups with significant differences in all-cause mortality over a 3-year follow-up period (**Table 2, Figure 4A**). The high-risk group exhibited a mortality rate nearly five-fold greater than the lower risk group (**Figure 4B**). This analysis was repeated for all-cause mortality over a 15-year follow-up period (**Online Figure 5**), and for CVD mortality over both a 3-year follow-up period (**Online Figure 6**) and 15-year follow-up period (**Online Figure 7**). As shown in **Online Figure 5**, the risk groups identified in **Figure 4** lose predictive value beyond approximately 5 years. The p-values obtained from a log-rank test comparing each pair of curves are provided in **Online Table 4**.

**Figure 4.**
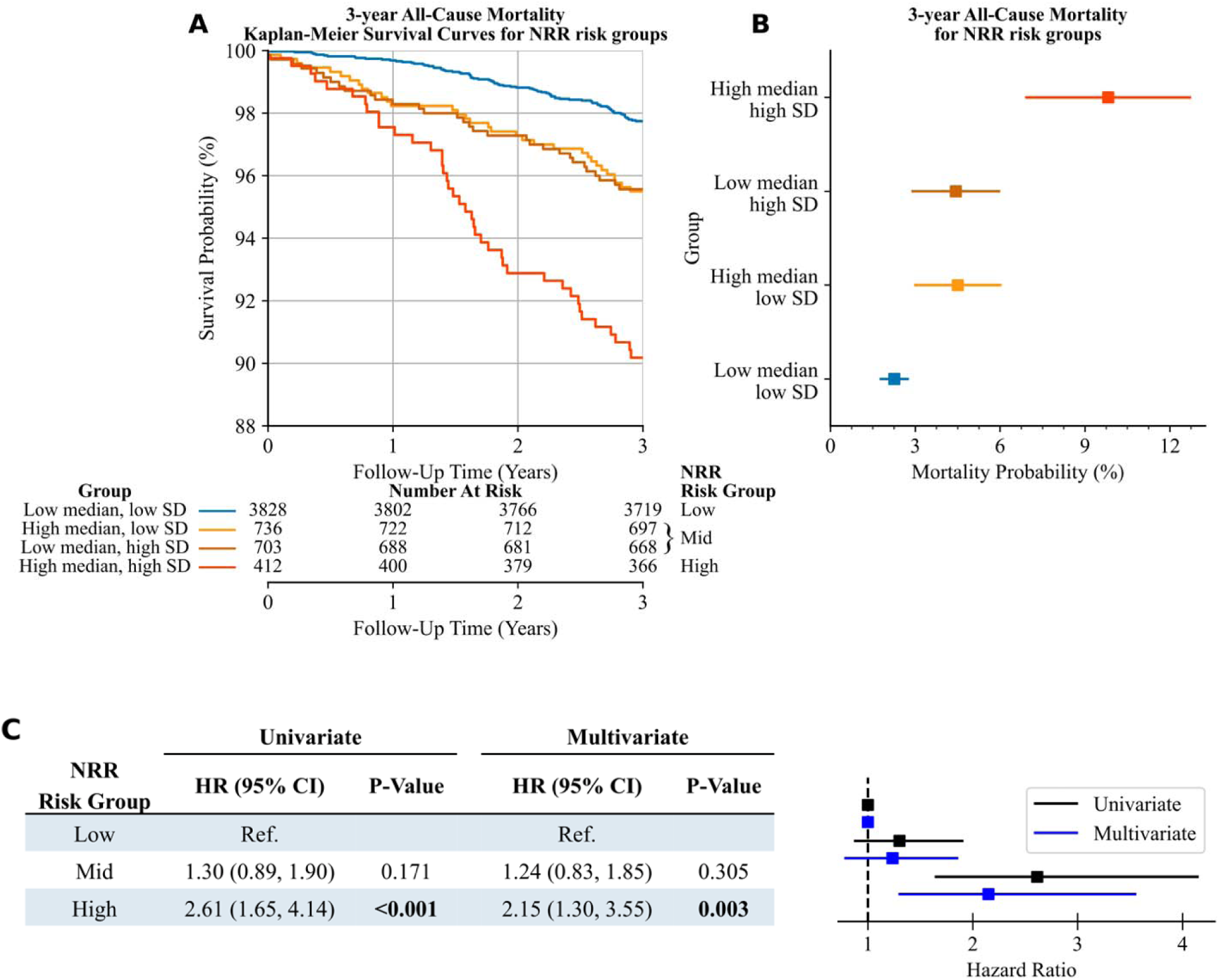
Survival analysis of risk groups stratified by the median and SD of NRR in relation to 3-year all-cause mortality. (A) 3-year all-cause mortality Kaplan-Meier survival curves for NRR risk groups. (B) 3-year cumulative incidence of all-cause death for NRR risk groups. (C) Hazard ratios for respiratory biomarker risk groups in relation to all-cause mortality, based on univariate and multivariate Cox proportional hazards regression models. Respiratory biomarker associations remain significant after adjusting for comorbidities. CI: confidence interval; HR: hazard ratio; NRR: nocturnal respiratory rate; SD: standard deviation.

### Cox Proportional-Hazard Analysis

Univariate Cox proportional-hazards analysis revealed significant associations between the NRR based high-risk group with 3-year all-cause mortality (Hazard Ratio: 2.61 (1.65, 4.14), p<0.001), but not for the mid-risk group (Hazard Ratio: 1.30 (0.89, 1.90), p=0.171). Being in the high-risk group remained a significant predictor for 3-year all-cause mortality after adjustment for age, gender, obesity (BMI>30), cardiovascular comorbidities such as heart failure, atrial fibrillation, diabetes and hypertension, and pulmonary comorbidities such as COPD, asthma, history of smoking, and severe sleep apnea (apnea-hypopnea index > 30) during the sleep study (Hazard Ratio: 2.15 (1.3, 3.55), p=0.003) (**Figure 4C**). Of note, the high-risk group was older, had a higher BMI, and had a greater burden of comorbidities compared to the low- and mid-risk groups. The hazard ratio and the corresponding p-value of all the variables used in the univariate and multivariate models for 3-year all-cause mortality are shown in **Online Table 5**.

## Discussion

We examined the relationship between nocturnal respiratory rate (NRR) dynamics and long-term mortality in a large, well-characterized cohort of clinically stable outpatients with cardiovascular disease undergoing elective polysomnography. Several key findings emerged. First, both cardiovascular and all-cause mortality exhibited a clear, dose-dependent relationship with median NRR. Individuals with higher nocturnal respiratory rates experienced substantially greater mortality risk, with patients exceeding 20 breaths per minute demonstrating markedly worse 3-year survival compared with those with lower rates. Second, within-night variability of NRR, quantified by the standard deviation, was independently associated with mortality. Third, and most notably, the combination of elevated baseline NRR and increased variability enabled substantially improved risk stratification. Individuals with both high median and high variability of NRR experienced approximately five-fold higher 3-year mortality compared with those with low values for both metrics, an association that persisted after adjustment for demographic characteristics and cardiopulmonary comorbidities. Together, these findings indicate that both the level and stability of nocturnal respiratory control carry prognostic information.

Interest in respiratory rate as a prognostic biomarker has grown alongside advances in wearable and noncontact sensing technologies that permit automated, longitudinal measurement with minimal patient burden^17,24–28^. Prior studies have demonstrated associations between elevated NRR and adverse outcomes in selected clinical contexts. For example, Dommasch et al. showed that increased nocturnal respiratory rate predicts non-sudden cardiac death in survivors of acute myocardial infarction, while Eick et al. reported that mean NRR during the first night of hospitalization predicts mortality in patients with acute coronary syndromes^8,13^. In community-based cohorts, Baumert et al. demonstrated that higher mean nocturnal respiratory rate is associated with increased long-term cardiovascular and all-cause mortality^12^. Our findings extend this body of work in several important ways. We focused specifically on clinically stable outpatients with established cardiovascular disease, leveraged waveform-level respiratory data rather than summary PSG outputs, and evaluated not only baseline respiratory rate but also within-night variability as a distinct physiological dimension.

The association between NRR variability and mortality is particularly noteworthy and may reflect instability in underlying cardiopulmonary or autonomic regulation. Increased respiratory variability has been observed preceding clinical deterioration and hospitalization in acute care settings and has been linked to impending ICU transfer^24,29,30^.In stable outpatients, elevated nocturnal respiratory variability may signal subclinical disease, impaired autonomic control, fluctuating pulmonary congestion, or altered chemoreflex sensitivity^14,15^. Importantly, the prognostic value of variability persisted after adjustment for heart failure, pulmonary disease, sleep apnea severity, and other comorbidities, suggesting that NRR variability captures physiological information not fully reflected by traditional diagnoses.

From a clinical perspective, these findings have several implications. First, nocturnal respiratory physiology appears to encode risk information that is not captured by conventional daytime measurements or single-point assessments. Second, longitudinal monitoring of NRR, particularly when both baseline level and variability are considered, may enable earlier identification of patients at elevated risk, potentially before overt clinical deterioration. This is especially relevant as respiratory rate during sleep is increasingly available through implanted devices, wearables, and noncontact home sensors. Third, our findings raise the possibility that NRR could be used to monitor the physiological impact of interventions that modify the underlying determinants of respiratory control, enabling assessment of whether such interventions alter risk trajectories over time.

### Limitations

Several limitations warrant consideration. The SHHS cohort is older and predominantly non-Hispanic, which may limit generalizability to younger or more diverse populations. Certain comorbidities, such as COPD, were relatively uncommon, and available comorbidity data lacked granularity regarding disease severity and treatment status. NRR metrics were derived from a single-night PSG, which may not fully capture night-to-night variability, although such measurement noise would be expected to bias results toward the null. Additionally, although PSGs were elective, some participants may have undergone testing during periods of transient illness or subclinical exacerbation. Our analysis focused on median and standard deviation of NRR; however, more complex temporal features or multivariate respiratory patterns may provide additional prognostic value. Finally, while NRR was strongly associated with mortality, it is unlikely to be causal, and mechanistic studies are needed to elucidate the biological pathways linking respiratory dynamics to adverse outcomes.

## Conclusions

In summary, we demonstrate that both the baseline level and within-night variability of nocturnal respiratory rate are independently associated with long-term cardiovascular and all-cause mortality in clinically stable outpatients with cardiovascular disease. Compared with prior studies, our work leveraged waveform-level respiratory analysis, focused on patients with established cardiovascular disease, included a substantial proportion of women, and identified NRR variability as a novel prognostic marker. By combining median NRR and its variability, we achieved clinically meaningful risk stratification beyond either metric alone. These findings underscore the potential of passive, longitudinal nocturnal respiratory monitoring as a scalable and interpretable tool for long-term risk assessment. As respiratory rate during sleep becomes increasingly accessible across clinical and consumer platforms, understanding its physiological determinants and clinical implications may open new avenues for early detection, monitoring, and intervention in cardiovascular disease.

## Data Availability

All data produced in the present study are available upon reasonable request to the authors.

## Acknowledgments

The Sleep Heart Health Study (SHHS) was supported by National Heart, Lung, and Blood Institute cooperative agreements U01HL53916 (University of California, Davis), U01HL53931 (New York University), U01HL53934 (University of Minnesota), U01HL53937 and U01HL64360 (Johns Hopkins University), U01HL53938 (University of Arizona), U01HL53940 (University of Washington), U01HL53941 (Boston University), and U01HL63463 (Case Western Reserve University). The National Sleep Research Resource was supported by the National Heart, Lung, and Blood Institute (R24 HL114473, 75N92019R002). J.E.O. was funded by NIH (K23HL151880). K.R.K. is funded Department of Defense HT94252410105, Wellcome Leap In Utero Program, NIH (NHLBI R33 HL168785, NHLBI R43HL157291, NIAID R43AI177239).

## Perspectives

### Competency in Medical Knowledge 1

Median nocturnal respiratory rate and its within-night variability independently predict all-cause and cardiovascular mortality at 3 years in a dose-dependent manner.

### Competency in Medical Knowledge 2

Combined assessment of median nocturnal respiratory rate and its variability enables stratification of clinically stable outpatients into low-, intermediate-, and high-risk groups for 3-year mortality.

### Translational Outlook 1

Because the association between nocturnal respiratory rate and mortality is unlikely to be causal, future studies should focus on identifying the physiological determinants of nocturnal respiratory rate and its variability, their dynamics, and their reversibility. As longitudinal respiratory monitoring becomes increasingly available through consumer and clinical devices, understanding these associations will be essential for translating respiratory biomarkers into clinical risk assessment and monitoring.

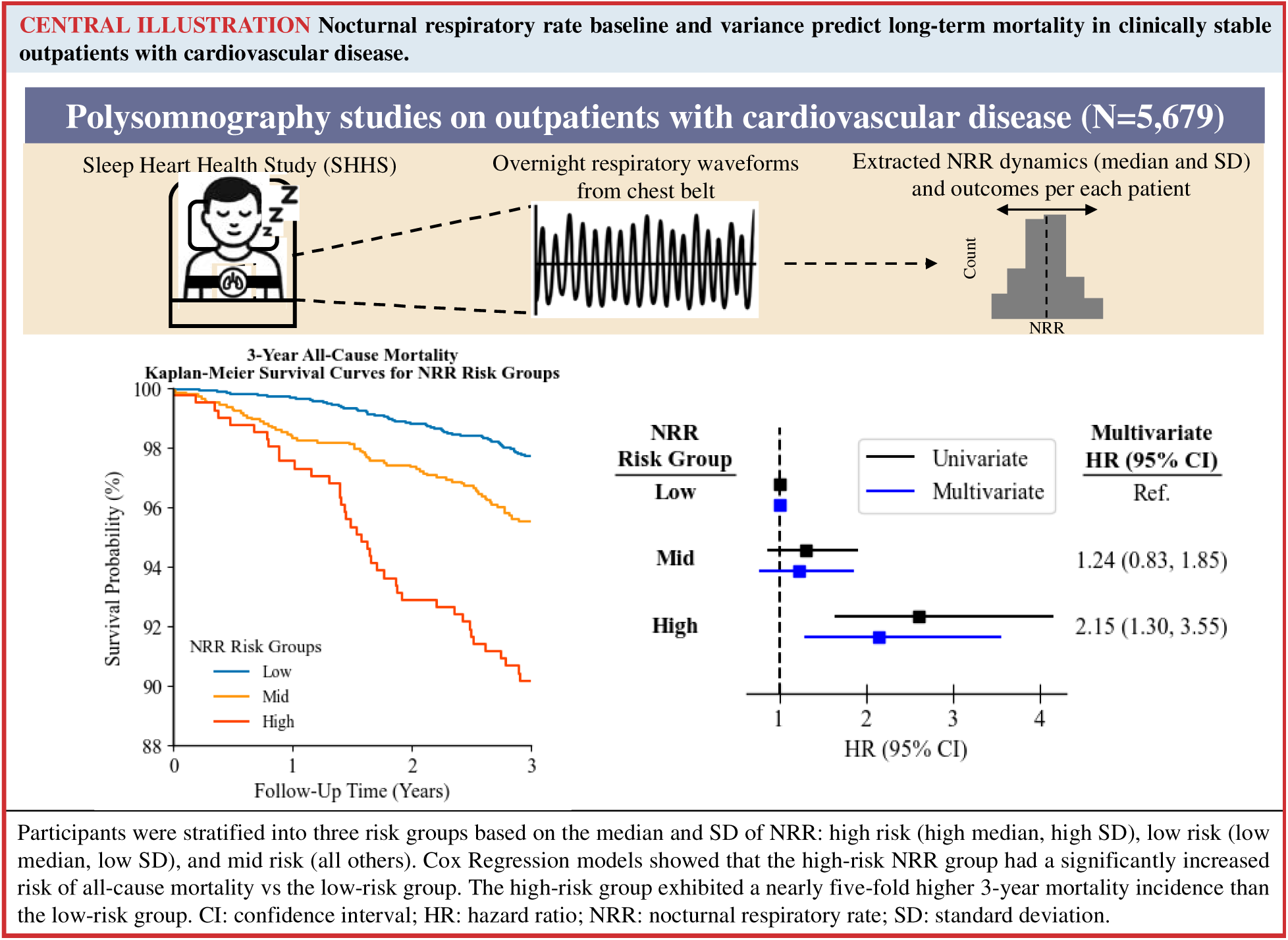
Central Illustration. Nocturnal respiratory rate baseline and variance predict long-term mortality in clinically stable outpatients with cardiovascular disease.

## Abbreviations

RR: Respiratory Rate
NRR: Nocturnal Respiratory Rate
brpm: Breaths per minute
SHHS: Sleep Health Heart Study

