## Supplemental Information for "Nocturnal Respiratory Rate and Variability Predict Long-term Mortality in Stable Outpatients with Cardiovascular Disease"

**Nocturnal respiratory biomarkers predict long-term mortality in outpatients with cardiovascular disease: Online Appendix**

Raimon Padrós-Valls, M.S.^a,b^

Keshav Gupta, M.B.B.S., M.S.^a^

Nick Harrington, Ph.D.^a^

Justin Yu, B.S.^a^

Jeremy E. Orr, M.D.^c^

Robert L. Owens, M.D.^c^

David Torres Barba, M.D., Ph.D.^d^

Kevin R. King, M.D., Ph.D.^a,d,*^

^a^Department of Bioengineering, Jacobs School of Engineering, University of California San Diego, La Jolla, CA, 92093, USA.

^b^Bioinformatics and Systems Biology, Jacobs School of Engineering, University of California San Diego, La Jolla, CA, 92093, USA.

^c^Division of Pulmonary, Critical Care and Sleep Medicine, Department of Medicine, University of California San Diego, La Jolla, CA, 92093USA

^d^Division of Cardiovascular Medicine, Department of Medicine, University of California San Diego, La Jolla, CA, 92093, USA.

**Online Figure 1. Example of respiratory signal recording over a single night.** brpm: breaths per minute; RR: respiratory rate.

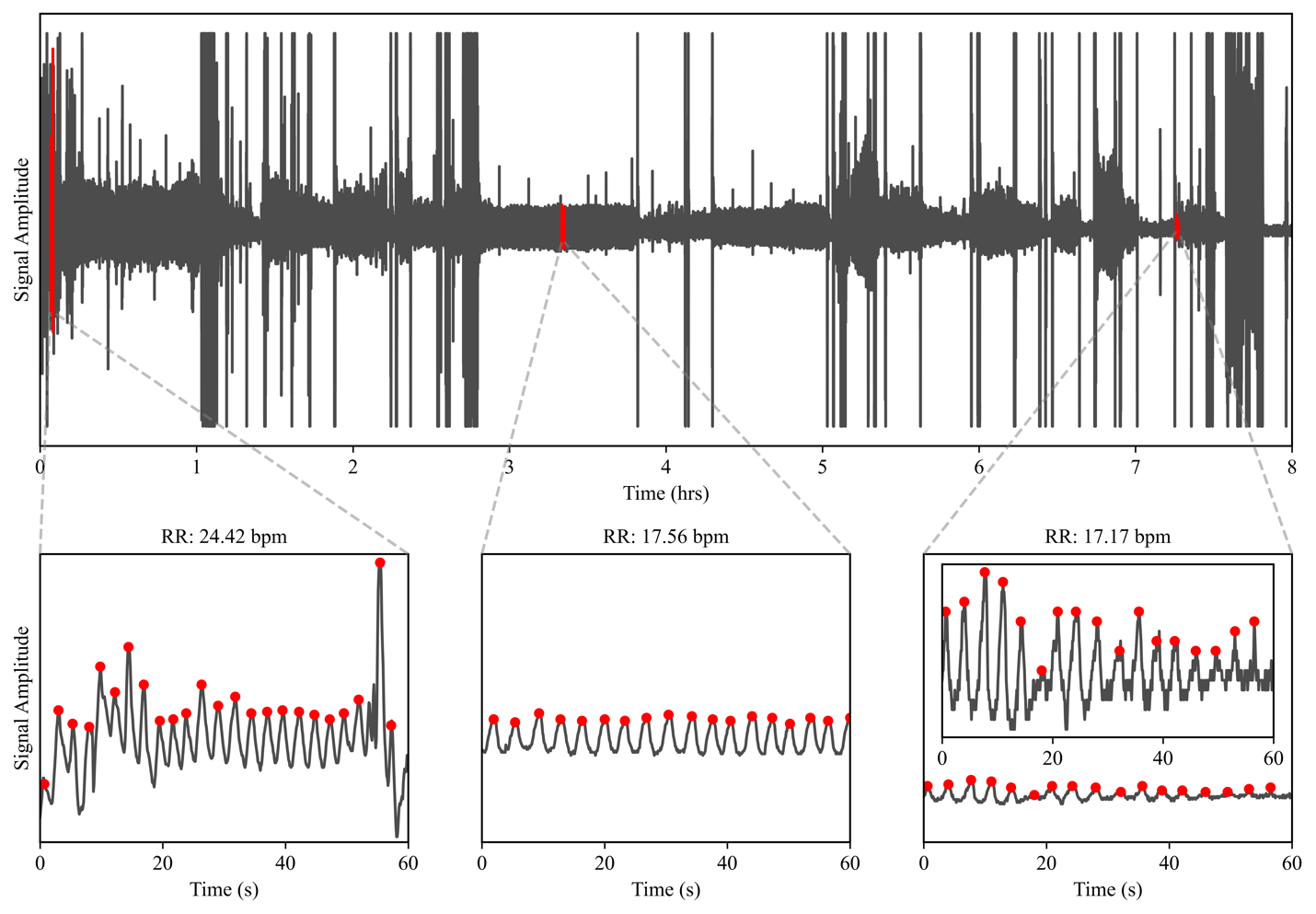

**Online Figure 2. 3-year CVD mortality analysis for bins of median NRR and bins of SD NRR.** Kaplan-Meier survival curves in a 3-year horizon (A) for median NRR bins, and (B) for SD NRR bins. 3-year cumulative incidence of death (C) for median NRR bins, and (D) for SD NRR bins. brpm: breaths per minute; CVD: cardiovascular; NRR: nocturnal respiratory rate; SD: standard deviation.

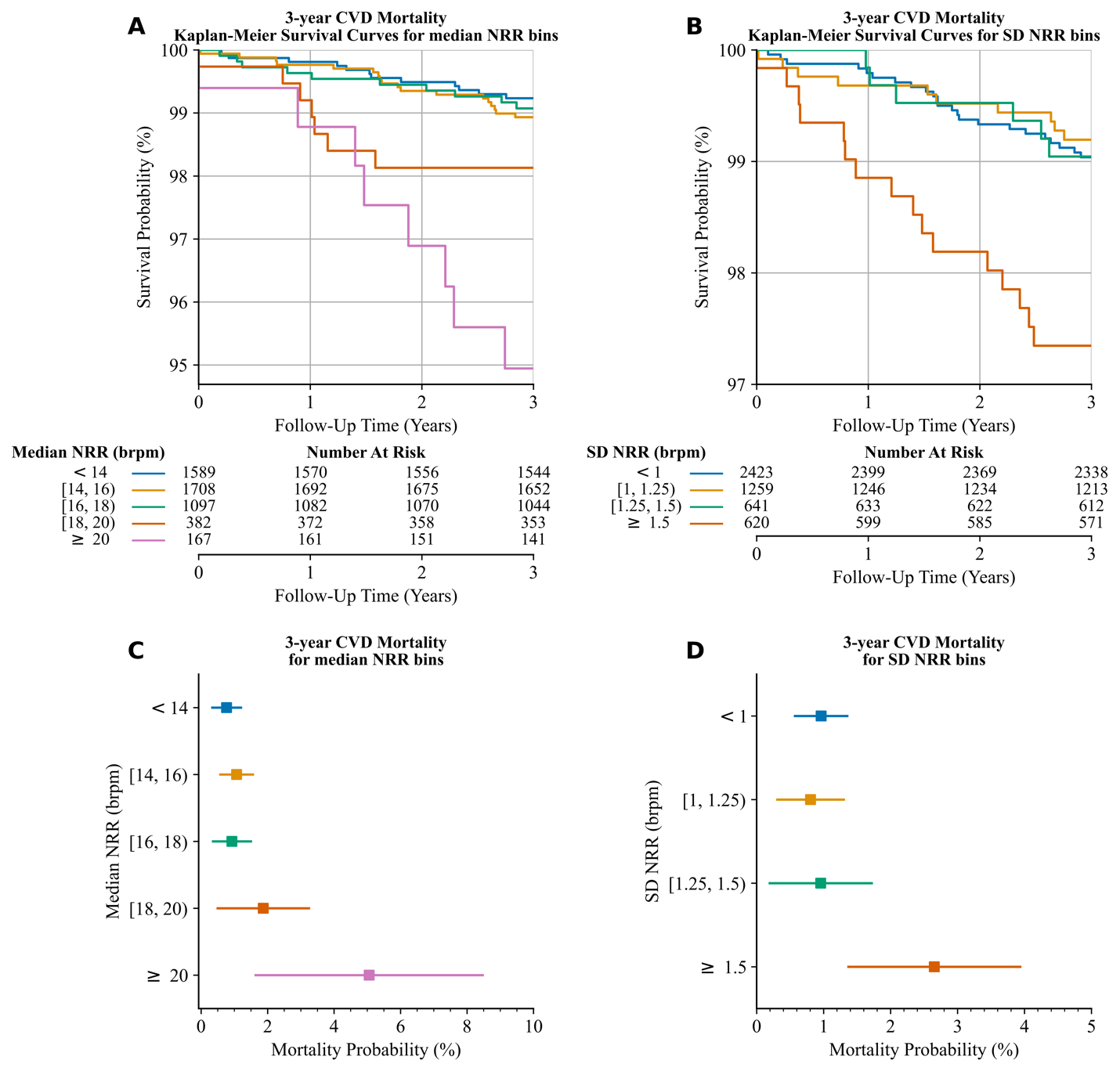

**Online Figure 3. 15-year all-cause mortality analysis for bins of median NRR and bins of SD NRR.** Kaplan-Meier survival curves in a 15-year horizon (A) for median NRR bins, and (B) for SD NRR bins. 15-year cumulative incidence of death (C) for median NRR bins, and (D) for SD NRR bins. brpm: breaths per minute; NRR: nocturnal respiratory rate; SD: standard deviation.

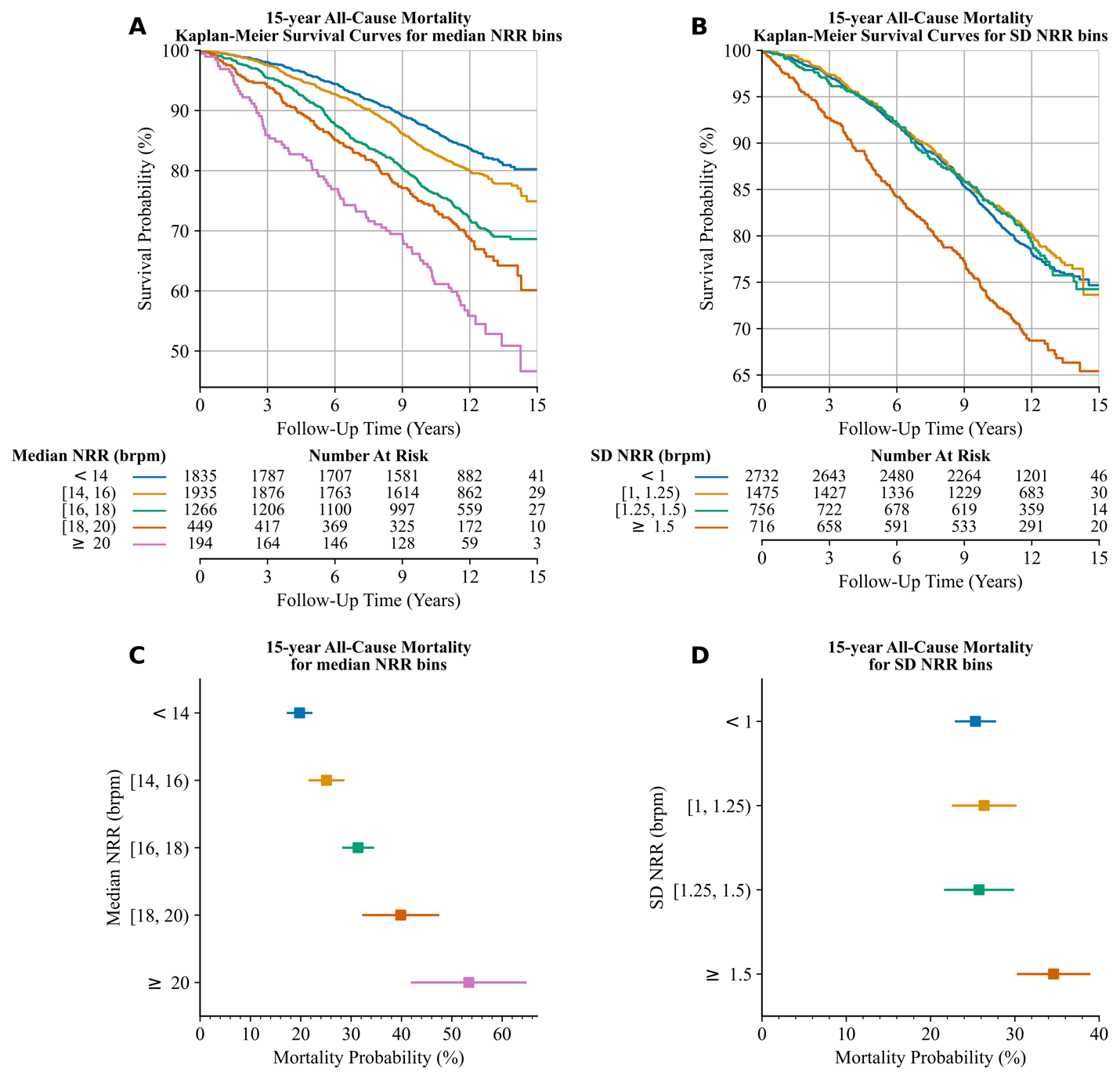

**Online Figure 4. 15-year CVD mortality analysis for bins of median NRR and bins of SD NRR.** Kaplan-Meier survival curves in a 15-year horizon (A) for median NRR bins, and (B) for SD NRR bins. 15-year cumulative incidence of death (C) for median NRR bins, and (D) for SD NRR bins. brpm: breaths per minute; CVD: cardiovascular; NRR: nocturnal respiratory rate; SD: standard deviation.

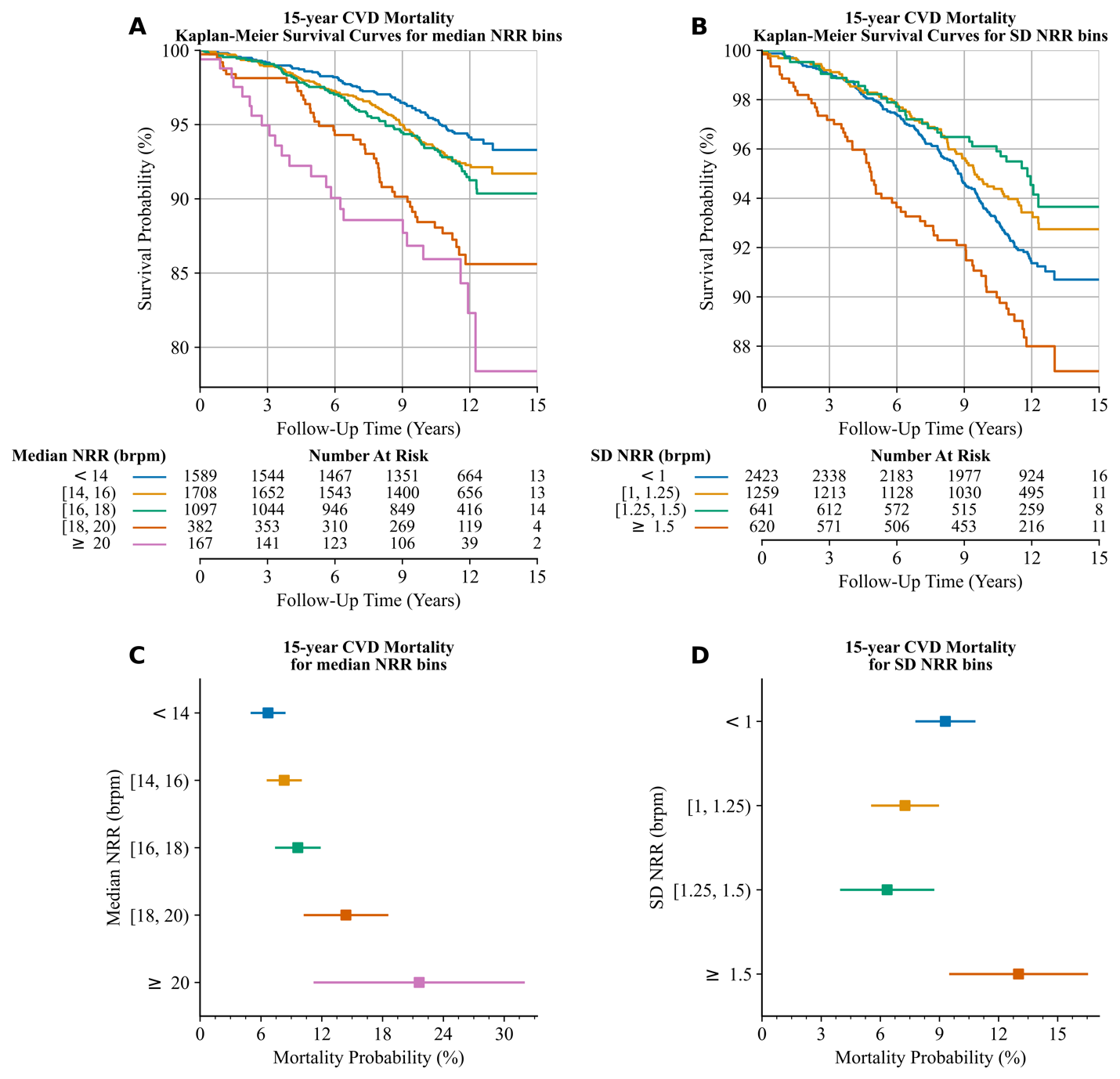

**Online Figure 5. Survival analysis of risk groups stratified by the median and SD of NRR in relation to 15-year all-cause mortality.** (A) 15-year all-cause mortality Kaplan-Meier survival curves for NRR risk groups. (B) 15-year cumulative incidence of all-cause death for NRR risk groups. NRR: nocturnal respiratory rate; SD: standard deviation.

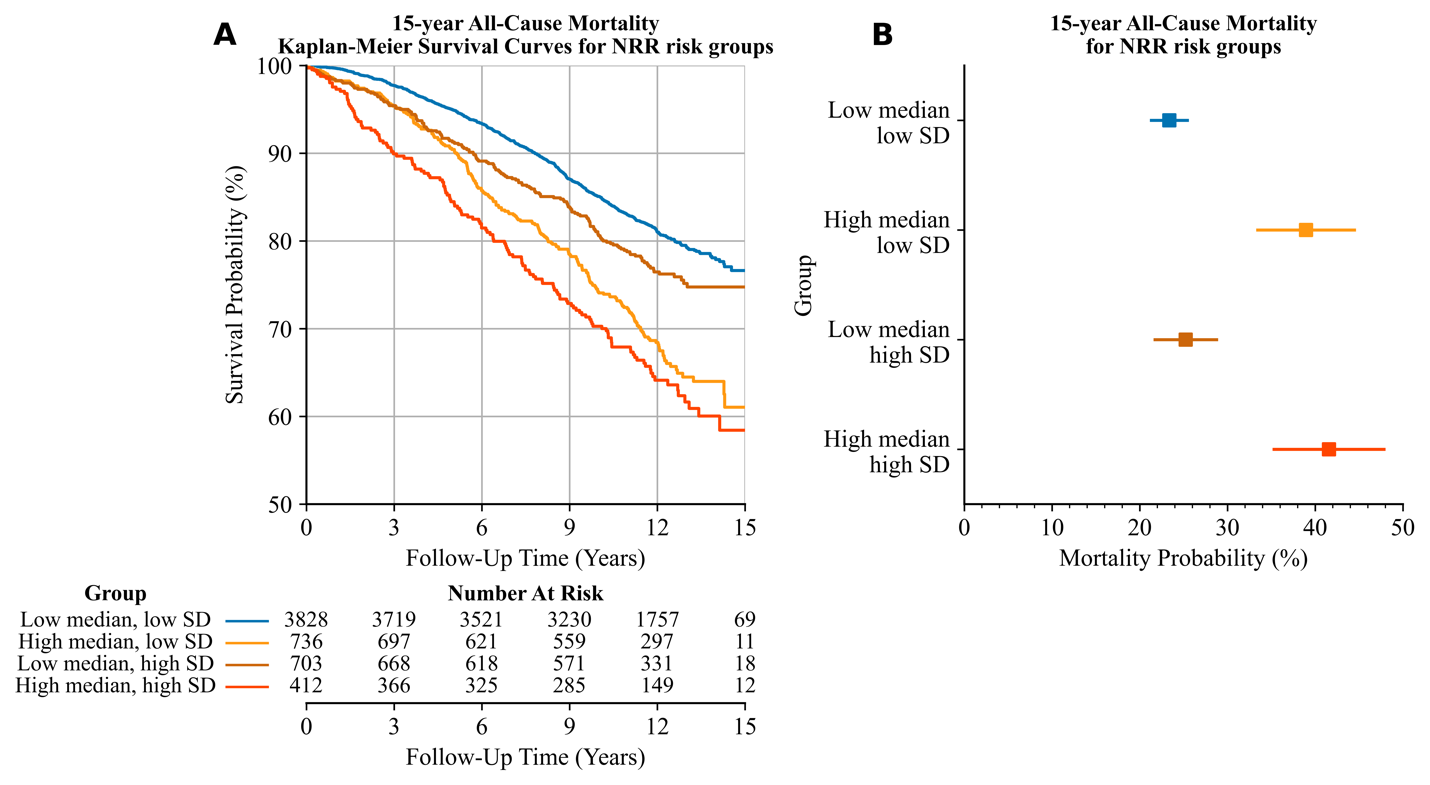

**Online Figure 6. Survival analysis of risk groups stratified by the median and SD of NRR in relation to 3-year CVD mortality.** (A) 3-year CVD mortality Kaplan-Meier survival curves for NRR risk groups. (B) 3-year cumulative incidence of CVD death for NRR risk groups. CVD: cardiovascular disease; NRR: nocturnal respiratory rate; SD: standard deviation.

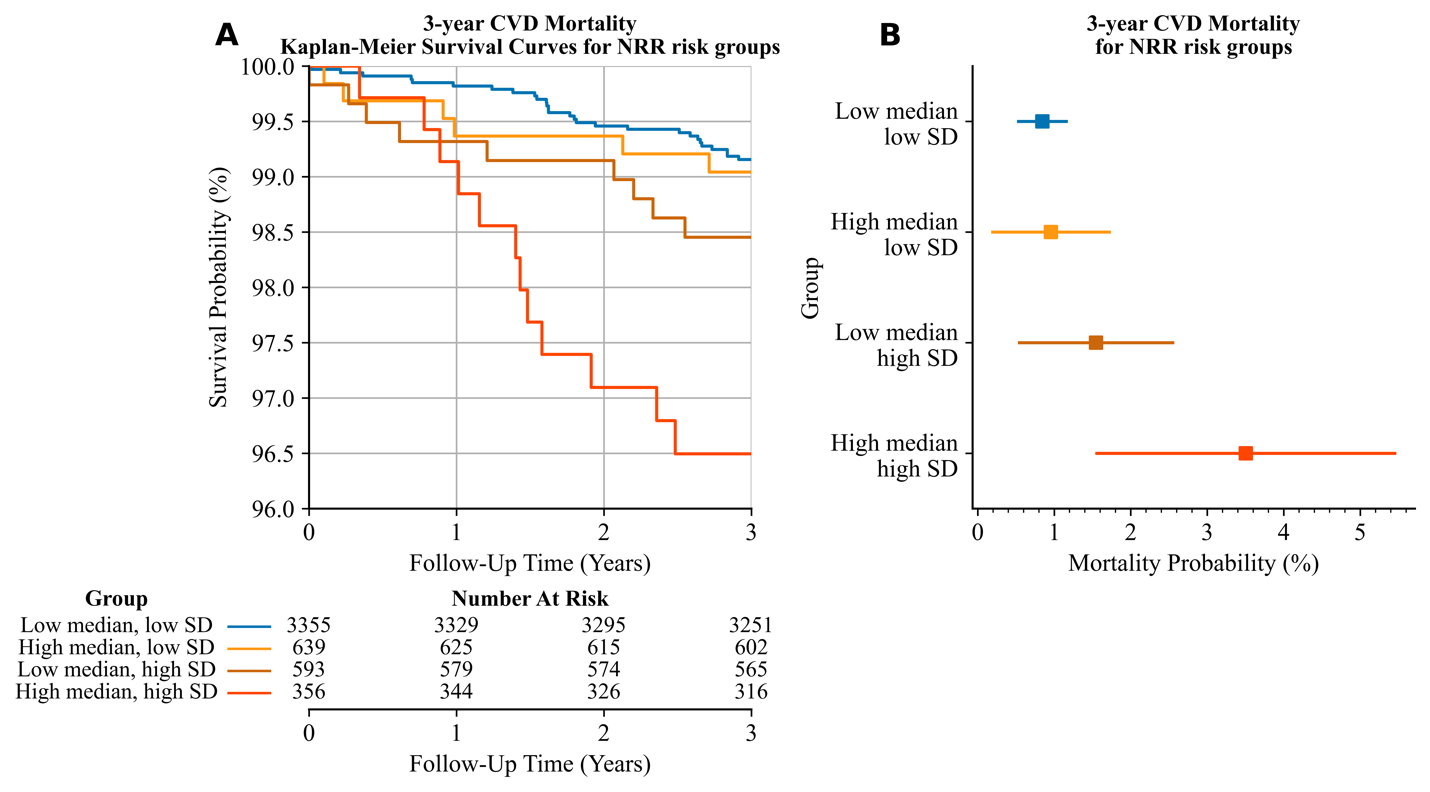

**Online Figure 7. Survival analysis of risk groups stratified by the median and SD of NRR in relation to 15-year CVD mortality.** (A) 15-year CVD mortality Kaplan-Meier survival curves for NRR risk groups. (B) 15-year cumulative incidence of CVD death for NRR risk groups. CVD: cardiovascular disease; NRR: nocturnal respiratory rate; SD: standard deviation.

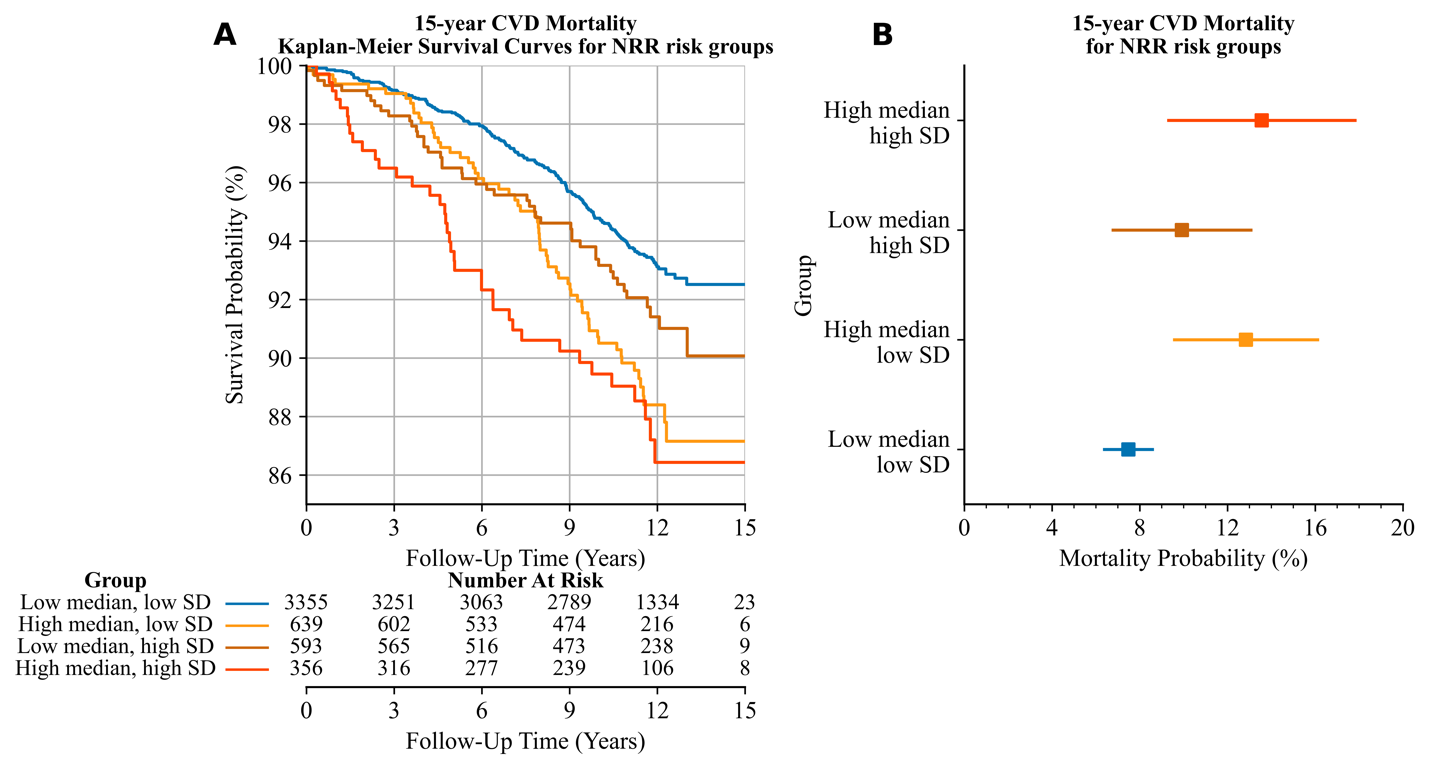

**Online Table 1. Respiratory rate epochs summary statistics.**

|  | **Mean** | **Median** | **SD** | **Total** |
| --- | --- | --- | --- | --- |
| Time available (hours) | 7.03 | 7.32 | 1.28 | 39,902.27 |
| Number of windows (count) | 301.57 | 319 | 76.71 | 1,712,656 |
| Number of valid windows (%) | 78.23 | 84.29 | 18.49 |  |
| Median NRR (brpm) | 15.22 | 15.02 | 2.37 |  |
| SD NRR (brpm) | 1.10 | 1.01 | 0.42 |  |

brpm: breaths per minute; NRR: nocturnal respiratory rate; SD: standard deviation.

**Online Table 2. Log-rank test p-values for median NRR Kaplan-Meier survival curves.** All pairs of curves from **Figure 3A**, **Online Figure 2A**, **Online Figure 3A**, and **Online Figure 4A** are compared using the pairwise log-rank test, while a multivariate log-rank test assesses overall differences.

| **Median NRR [brpm] Survival Curve** | |  | **P-Value** | | | |
| --- | --- | --- | --- | --- | --- | --- |
| **1** | **2** |  | **3-Years All-Cause Mortality** | **3-Years CVD Mortality** | **15-Years All-Cause Mortality** | **15-Years CVD Mortality** |
| < 14 | [14, 16) |  | 0.325 | 0.370 | **0.004** | **0.043** |
| < 14 | [16, 18) |  | **<0.001** | 0.652 | **<0.001** | 0.011 |
| < 14 | [18, 20) |  | **<0.001** | **0.047** | **<0.001** | **<0.001** |
| < 14 | >= 20 |  | **<0.001** | **<0.001** | **<0.001** | **<0.001** |
| [14, 16) | [16, 18) |  | **<0.001** | 0.723 | **0.004** | 0.464 |
| [14, 16) | [18, 20) |  | **<0.001** | 0.186 | **<0.001** | **<0.001** |
| [14, 16) | >= 20 |  | **<0.001** | **<0.001** | **<0.001** | **<0.001** |
| [16, 18) | [18, 20) |  | 0.250 | 0.137 | 0.090 | **0.005** |
| [16, 18) | >= 20 |  | **<0.001** | **<0.001** | **<0.001** | **<0.001** |
| [18, 20) | >= 20 |  | **0.001** | **0.049** | **0.002** | 0.266 |
| Multivariate | |  | **<0.001** | **<0.001** | **<0.001** | **<0.001** |

brpm: breaths per minute; CVD: cardiovascular death; NRR: nocturnal respiratory rate.

**Online Table 3. Log-rank test p-values for NRR standard deviation Kaplan-Meier survival curves.** All pairs of curves from **Figure 3B**, **Online Figure 2B**, **Online Figure 3B**, and **Online Figure 4B** are compared using the pairwise log-rank test, while a multivariate log-rank test assesses overall differences.

| **SD NRR [brpm] Survival Curve** | |  | **P-Value** | | | |
| --- | --- | --- | --- | --- | --- | --- |
| **1** | **2** |  | **3-Years All-Cause Mortality** | **3-Years CVD Mortality** | **15-Years All-Cause Mortality** | **15-Years CVD Mortality** |
| < 1 | [1, 1.25) |  | 0.746 | 0.638 | 0.320 | 0.084 |
| < 1 | [1.25, 1.5) |  | 0.307 | 0.987 | 0.735 | **0.023** |
| < 1 | >=1.5 |  | **<0.001** | **<0.001** | **<0.001** | **0.007** |
| [1, 1.25) | [1.25, 1.5) |  | 0.242 | 0.742 | 0.691 | 0.345 |
| [1, 1.25) | >=1.5 |  | **<0.001** | **0.001** | **<0.001** | **<0.001** |
| [1.25, 1.5) | >=1.5 |  | **0.001** | **0.023** | **<0.001** | **<0.001** |
| Multivariate | |  | **<0.001** | **0.002** | **<0.001** | **<0.001** |

brpm: breaths per minute; CVD: cardiovascular death; NRR: nocturnal respiratory rate; SD: standard deviation.

**Online Table 4. Log-rank test p-values for NRR risk groups.** All pairs of curves from **Figure 4A**, **Online Figure 5**, **Online Figure 6A**, and **Online Figure 7A** are compared using the pairwise log-rank test, while a multivariate log-rank test assesses overall differences.

| **Risk Group Survival Curve** | |  | **P-Value** | | | |
| --- | --- | --- | --- | --- | --- | --- |
| **1** | **2** |  | **3-Year All-Cause Mortality** | **15-Year All-Cause Mortality** | **3-Year CVD Mortality** | **15-Year CVD Mortality** |
| Low Median, Low SD | High Media, Low SD |  | **<0.001** | **<0.001** | 0.766 | **<0.001** |
| Low Median, Low SD | Low Median, High SD |  | **<0.001** | **0.011** | 0.104 | 0.095 |
| Low Median, Low SD | High Median, High SD |  | **<0.001** | **<0.001** | **<0.001** | **<0.001** |
| High Median, Low SD | Low Median, High SD |  | 0.955 | **0.001** | 0.356 | 0.136 |
| High Median, Low SD | High Median, High SD |  | **<0.001** | 0.137 | **0.005** | 0.432 |
| Low Median, High SD | High Median, High SD |  | **<0.001** | **<0.001** | 0.054 | **0.047** |
| Multivariate | |  | **<0.001** | **<0.001** | **<0.001** | **<0.001** |

CVD: cardiovascular disease; SD: standard deviation.

**Online Table 5. Association of covariates with all-cause mortality during a 3-year follow-up using a univariate and multivariate Cox Hazards Regression model.** The multivariate model adjusts for NRR risk groups, age, obesity, sex, and comorbidities including diagnosis of heart failure, atrial fibrillation, hypertension, COPD, and asthma, smoking history and severe sleep apnea at baseline.

|  | **Univariate Model** | |  | **Multivariate Model** | |
| --- | --- | --- | --- | --- | --- |
|  | **HR (95% CI)** | **P-Value** |  | **HR (95% CI)** | **P-Value** |
| NRR Risk Group |  |  |  |  |  |
| Low | Ref. |  |  | Ref. |  |
| Mid | 1.30 (0.89-1.9) | 0.171 |  | 1.24 (0.83-1.85) | 0.305 |
| High | 2.61 (1.65-4.14) | <0.001 |  | 2.15 (1.3-3.55) | 0.003 |
| Age (per 5 years) | 1.72 (1.54-1.93) | <0.001 |  | 1.65 (1.47-1.85) | <0.001 |
| Obesity (BMI>30) | 0.65 (0.42-1.0) | 0.051 |  | 0.78 (0.5-1.22) | 0.273 |
| Female | 0.7 (0.49-0.99) | 0.044 |  | 0.7 (0.49-1.02) | 0.063 |
| Heart Failure | 6.37 (3.71-10.92) | <0.001 |  | 2.77 (1.54-4.98) | 0.001 |
| Atrial Fibrillation | 3.48 (1.7-7.12) | 0.001 |  | 1.16 (0.55-2.47) | 0.696 |
| Diabetes | 2.89 (1.88-4.45) | <0.001 |  | 1.68 (1.08-2.62) | 0.022 |
| Hypertension | 2.01 (1.41-2.87) | <0.001 |  | 0.96 (0.65-1.4) | 0.818 |
| COPD | 1.81 (0.58-5.7) | 0.308 |  | 1.06 (0.32-3.51) | 0.929 |
| Smoke | 1.12 (0.79-1.59) | 0.523 |  | 1.07 (0.75-1.54) | 0.711 |
| Asthma | 1.24 (0.71-2.16) | 0.446 |  | 1.58 (0.88-2.84) | 0.126 |
| Severe Sleep Apnea (AHI > 30) | 1.46 (0.77-2.79) | 0.248 |  | 0.97 (0.5-1.89) | 0.924 |

AHI: apnea-hypopnea index; BMI: body mass index; CI: confidence interval; COPD: chronic obstructive pulmonary disease; HR: hazard ratio; NRR: nocturnal respiratory rate.
